# Epidemiological and Programmatic Insights into Progress toward Ending AIDS by 2030: Lessons from Viet Nam’s HIV Response

**DOI:** 10.64898/2026.07.01.26357006

**Authors:** Pham Duc Manh, Machiko Otani, Van Thi Thuy Nguyen, Phan Thi Thu Huong, Bui Hoang Duc, Fukushi Morishita, Nguyen Thi Minh Tam, Doan Thi Thuy Linh, Do Thi Nhan, Rajendra Prasad Hubraj Yadav, Kiyohiko Izumi

## Abstract

**Introduction:** Viet Nam has achieved one of the fastest declines in new HIV infections in the WHO Western Pacific Region. In 2020, the Government launched the National Strategy to End the AIDS Epidemic by 2030. At its midpoint in 2025, a national epidemiological and programmatic review was undertaken to assess the country’s progress and remaining challenges.

**Methods:** We conducted a review of the national HIV surveillance and programme data and the Joint United Nations Programme on HIV/AIDS database, focusing on four components: disease burden, prevention, testing, and treatment programmes.

**Results:** Estimated annual new HIV infections among adults declined from 14,000 (2010) to 6,117 (2024), a 57% reduction, while the number of PLHIV stabilised at 267,455 in 2024. The epidemic has shifted from being mainly among people who inject drugs (PWID) in the 1990s and early 2000s to being increasingly concentrated among men who have sex with men (MSM) and transgender people (TG), who accounted for 58% of new infections in 2024. In 2024, 88% of people living with HIV (PLHIV) knew their status, 79% of those diagnosed received antiretroviral therapy (ART), and 96% of people on ART achieved viral suppression. Harm reduction outcomes were strong, with opioid substitution therapy (OST) coverage around 40% since 2020 and safe injecting practices among PWID above 90% over the last ten years. The pre-exposure prophylaxis programme expanded rapidly in recent years, reaching 17.4% of the MSM. HIV testing volume was 3.4 million in 2024, with an increasing number of confirmatory testing laboratories. HIV status awareness in 2024 was below the 80% national target for sex workers (59.0%), PWID (62.5%), and MSM (79.2%), but exceeded the target for TG (94.1%). The high rate of viral load suppression indicates strong adherence and reflects the overall quality of treatment services.

**Conclusions:** Viet Nam’s progress reflects two decades of sustained harm reduction among people who inject drugs, early adoption of community-led and decentralised HIV testing innovations, rapid scale-up of PrEP, and consistently high viral suppression among people on ART. These combined system and community-based approaches offer transferable lessons for other low- and middle-income countries.

## Introduction

The human immunodeficiency virus (HIV) epidemic has been a significant public health challenge in Viet Nam, impacting economic growth and development[1]. The country has transformed its HIV response through strong national leadership, community engagement and integration of services into the broader health system[2]. In 2010, the Government demonstrated strong leadership by launching the first National Strategy for 2010-2020 with a long-term vision to 2030. Building on this foundation, the current National Strategy for 2021-2030[3] reinforces political commitment, aiming to reduce new HIV infections, HIV-related deaths, and the socio-economic burden of the epidemic with a goal to End the AIDS Epidemic by 2030. The National Strategy priorities for 2021-2030 include community-led and stigma-free prevention services for key populations, achieving 95-95-95 targets and sustainable financing.

Viet Nam has achieved one of the most rapid and substantial declines in new HIV infections in the WHO Western Pacific Region[4]. At the same time, Viet Nam has sustained progress toward the 95–95–95 targets, achieving 88% of people living with HIV aware of their status, 79% of those diagnosed receiving antiretroviral therapy, and 96% of those on treatment who are virally suppressed in 2024[5]. At the same time, 48% of the total financial resources allocated for HIV responses in 2021-2024 were from international donors, raising concerns about sustainability and service continuity.

In 2025, the country is at the mid-point of the National Strategy (2021-2030). We conducted an epidemiological and programmatic review in order to evaluate the country’s progress, identify the key drivers of its success, and highlight the remaining challenges to achieve Ending the AIDS Epidemic by 2030.

## Methods

We conducted a descriptive analysis of national and subnational data to assess epidemiological trends and program implementation as a part of the midterm programme review (2021-2024) for the implementation of the National Strategy to End AIDS Epidemic by 2030 (2021-2030). The results of program performance will inform the implementation of the National Strategy toward 2030.

### Review framework and data sources

The populations of interest were people living with HIV (PLHIV) and the following key populations (KPs) at increased risk of acquiring HIV: men who have sex with men (MSM), people who inject drugs (PWID), female sex workers (FSW), and transgender people (TG)[6].

We reviewed the following four components: A. Disease burden, B. Prevention programme, C. Testing programme, and D. Treatment programme. The HIV disease burden (A) was assessed using modelled data from the AIDS Epidemic Model [7], national HIV case reporting and data on KPs from sentinel surveillance and behavioural surveys. Programmatic review (B-D) was conducted using national HIV programme reports and monitoring data, which were complemented by data on KPs from sentinel surveillance and behavioural surveys. Reviewed indicators were informed by both the WHO strategic information guideline [8] and the monitoring and evaluation framework in the National Strategy to End the AIDS Epidemic by 2030 of Viet Nam [3]. Disease burden focused on new HIV infections, HIV-related deaths, prevalence and demographics of PLHIV. Program components were assessed in terms of outputs, coverage, and the impact of interventions. Where applicable, trends were analysed for the periods with available data.

### Ethics

Ethical approval was unnecessary for this study because we only used data collected as part of surveillance activities or aggregated programme data, and no individual-level data were obtained.

## Results

### A. Disease burden

In 2024, the estimated number of adults aged 15 or over living with HIV reached 267,455, reflecting only a modest annual increase since exceeding 260,000 in 2020. The annual estimated number of new HIV infections among adults was 6,177, which declined by 57% (2010–2024) and 34% (2020–2024) —equivalent to annualised compound reductions of 5.7% and 9.8%, respectively. The 57% reduction since 2010 ranks among the fastest declines in the WHO Western Pacific Region (Supplementary Figure S1). Notably, within the current National Strategy cycle, new infections dropped sharply by 13% between 2023 and 2024 [9]. The percentage change per year shows a steady decline after 2015 (Figure 1). [4] Estimated HIV-related mortality among adults has plateaued at 5,600-5,900 deaths annually since 2020, following a gradual rise from 5,089 in 2015 to 5,878 in 2019[9]. The estimated number of PLHIV, new infections, and number of HIV-related deaths among children aged between 0 and 14 years was 3358, 168 and 108, respectively.

**Figure 1.**
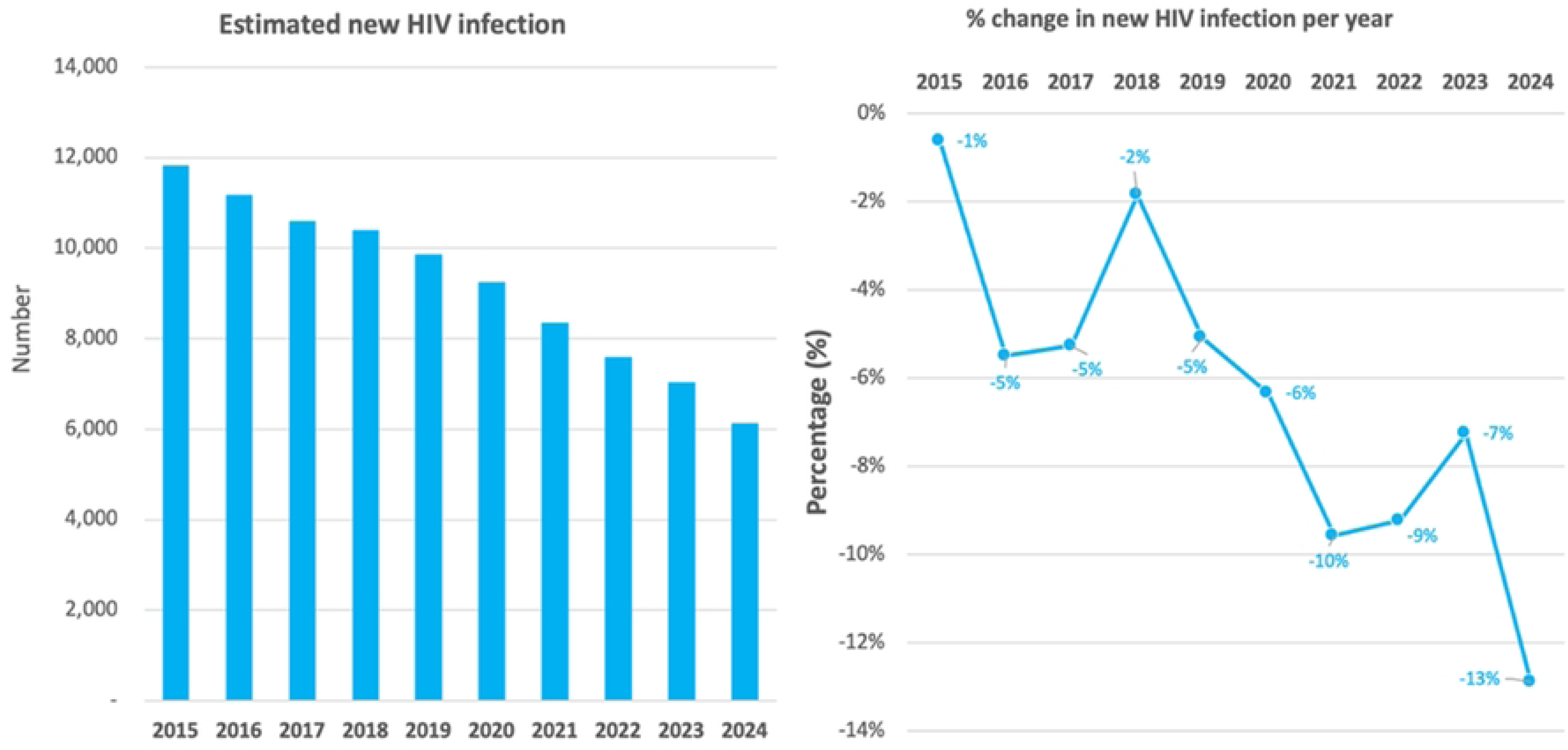
Estimated new HIV infections in Viet Nam, 2015-2024.

The estimated distribution of populations in new HIV infections (Figure 2) highlights a marked shift in affected populations: initially concentrated among PWID, the epidemic is now increasingly concentrated among MSM and TG, who together accounted for 58% of new infections in 2024[9]. HIV prevalence remains highest among PWID (9.0%, 2023) and MSM (7.2%, 2024), with lower rates among female sex workers (FSW) (2.0%, 2024)[10]. In comparison, prevalence among adults aged between 15 and 49 is 0.4%[4]. The vast majority of HIV reported cases were among men (83% in 2024) and in people aged between 16 and 39 years (69% in 2024)[11].

**Figure 2.**
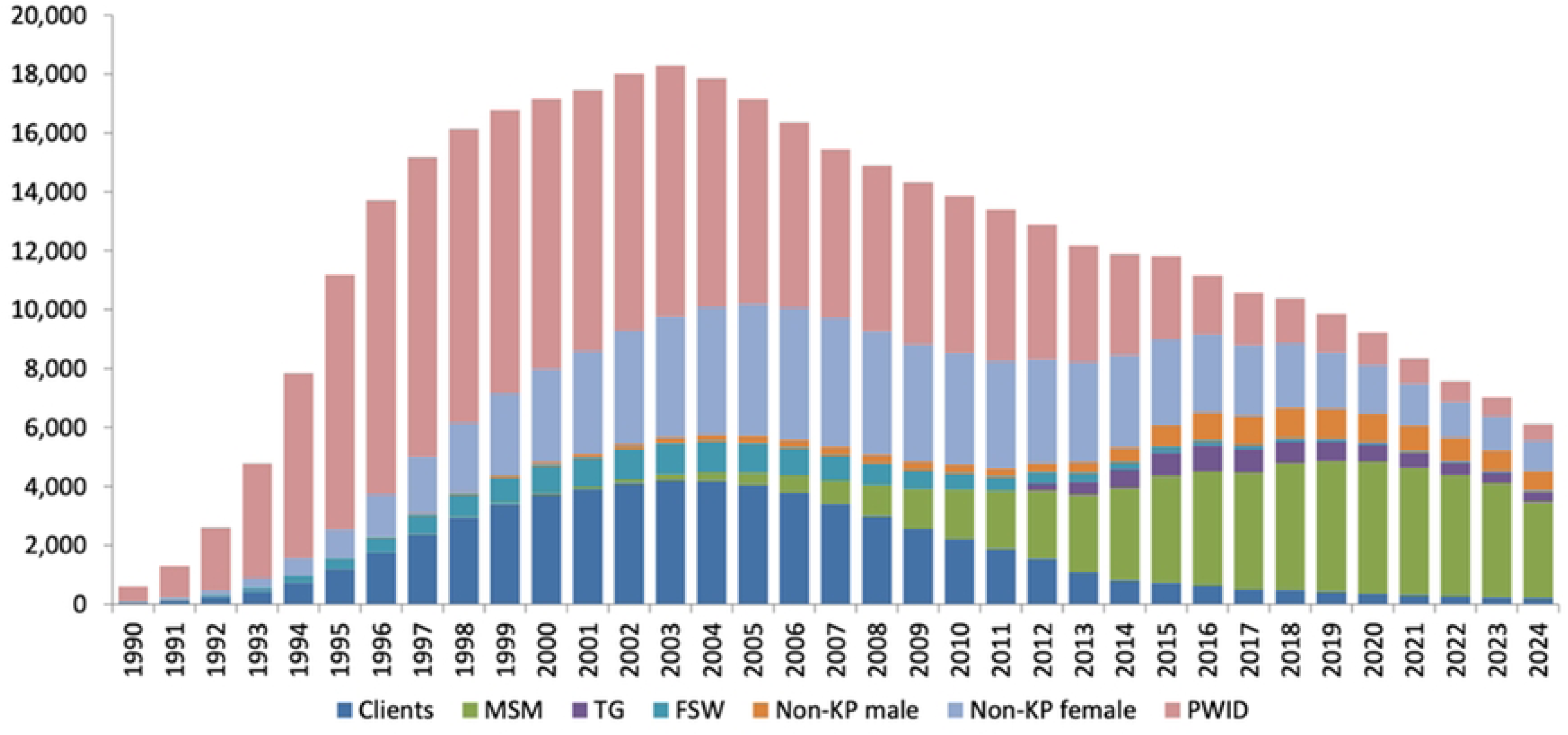
Estimated number of new HIV infections and population distribution, 1990-2024. FSW: female sex workers, KP: key population, MSM: men who have sex with men, PWID: people who inject drugs, TG: transgender people.

### B. Prevention programme

Since 2020, the proportion of PWID receiving opioid substitution therapy (OST) has remained at around 40% [11], achieving the national target of 40% and being close to the global target of 50% [12] in recent years. Needle and syringe programs have also achieved notable success, with distribution per PWID remaining above 100 units annually for over a decade until 2020, except for a brief decline between 2013 and 2015 [13]. Remarkably, safe injecting practices, defined as using sterile injecting equipment the last time they injected drugs, among PWID have remained consistently high, exceeding 90% throughout 2011–2023[13] (Figure 3).

**Figure 3.**
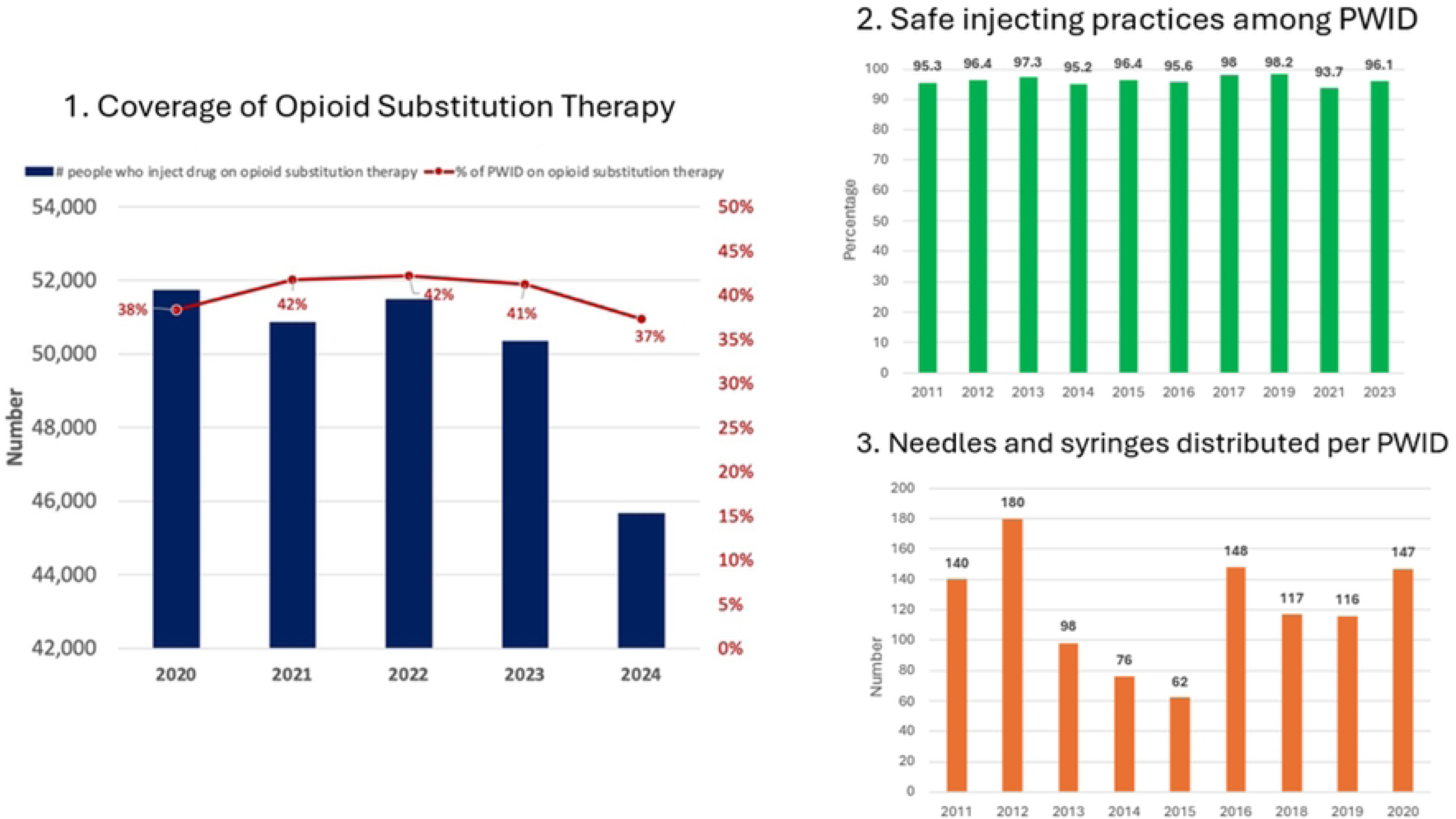
Harm reduction programme data in Viet Nam. PWID: people who inject drugs

At the same time, condom promotion among other key populations, FSW and MSM, has been highly successful. In 2022, prevention programs distributed an average of 299 condoms per FSW and 72 per MSM [13]. In 2024, 74.6% of FSW and 90.5% of MSM reported using a condom during their last sex with a non-marital, non-cohabiting partner [13] (Figure 4). Notably, condom use among FSW has consistently remained at a very high level, sustaining close to the global target of 90% since 2011[12].

**Figure 4.**
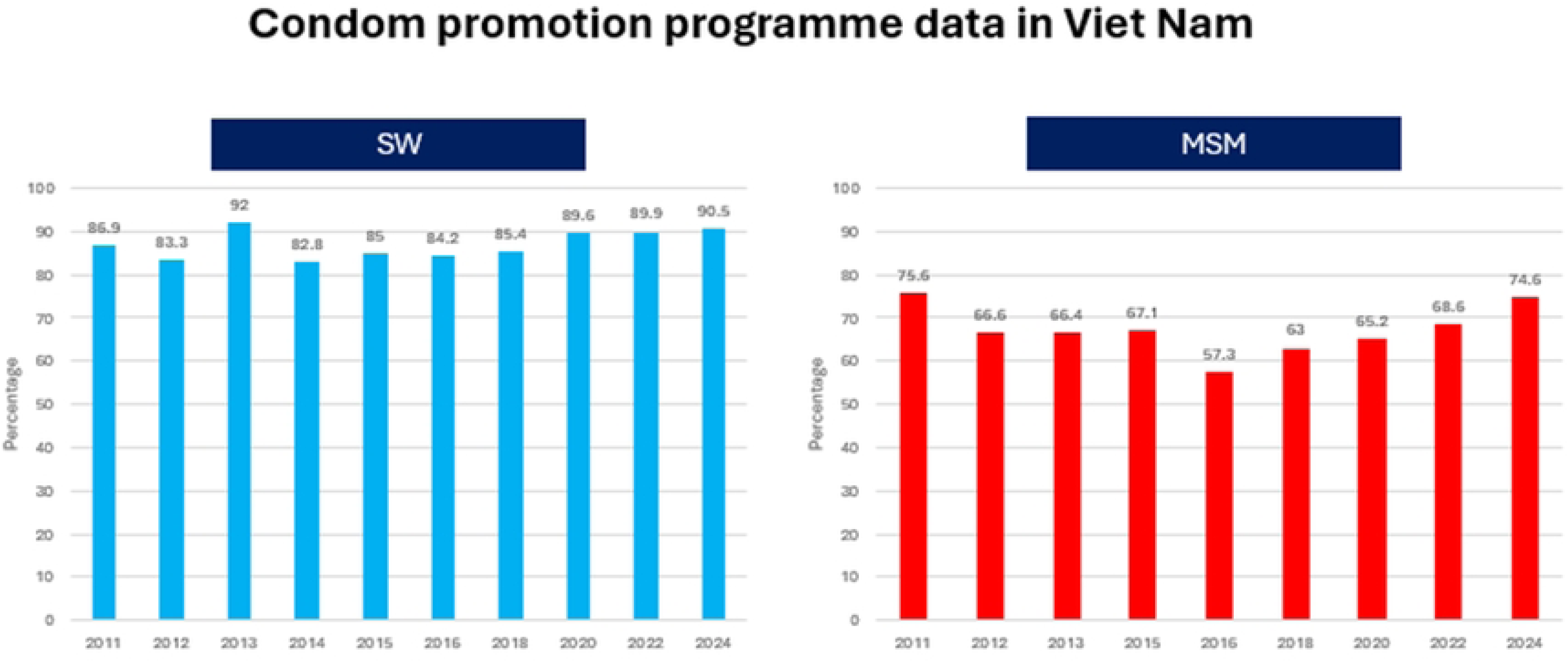
Condom promotion programme data in Viet Nam. MSM: men who have sex with men, SW: sex workers.

In addition, the pre-exposure prophylaxis (PrEP) programme expanded rapidly over the years (Figure 5). The number of new clients, cumulative number of clients received PrEP at least once, clients on PrEP in respective year, and PrEP continuation for at least three months steadily increased over the last five years[14]. As of 2024, 74,890 individuals had used PrEP at least once. However, based on the estimated MSM population size (429,419 in 2024 [9]), coverage remains at only 17.4%, falling short of the national target of 30% by 2025.

**Figure 5.**
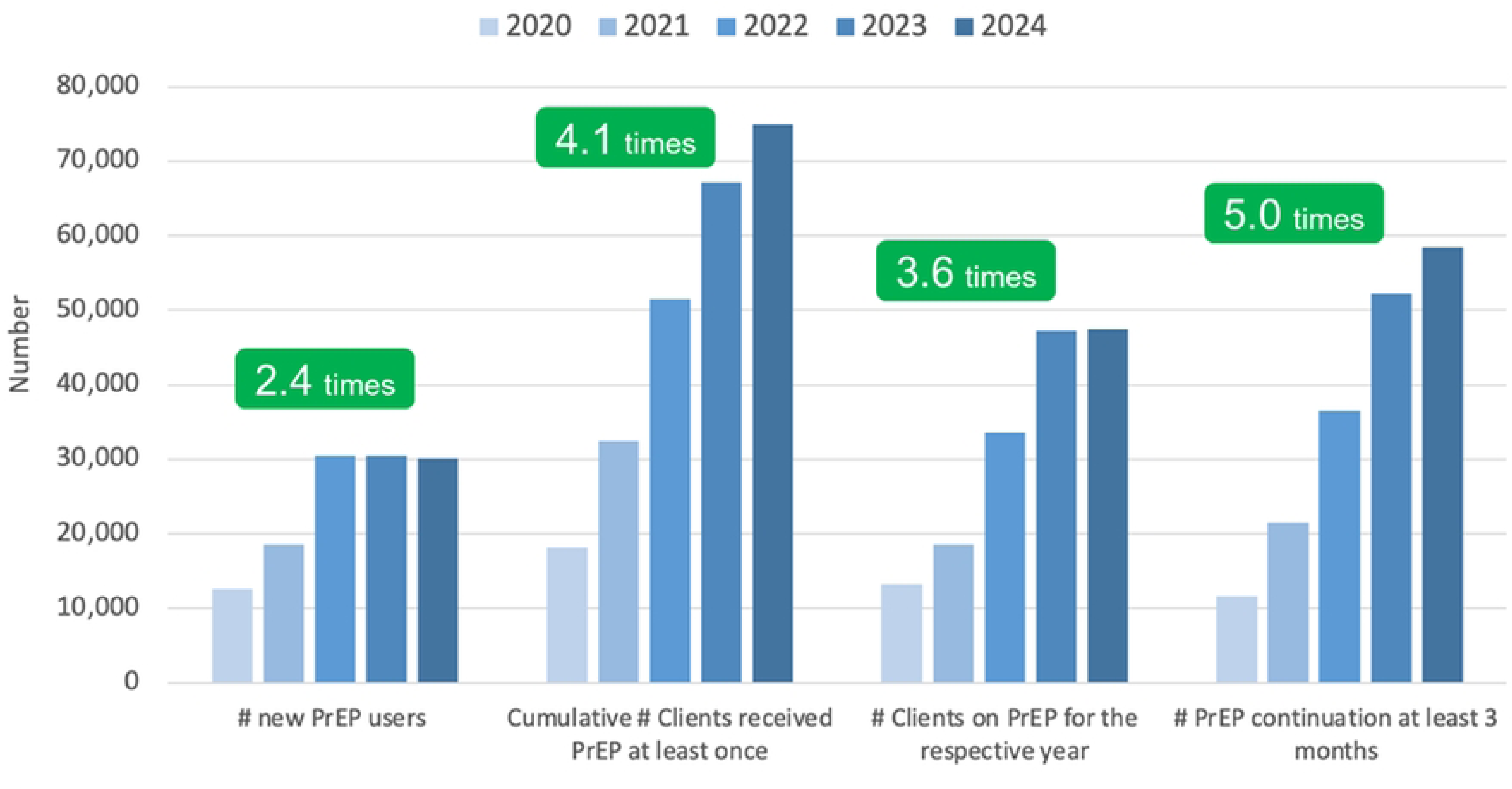
Pre-exposure prophylaxis (PrEP) programme data, 2020-2024. #: Number Comparisons are based on figures in 2020 and 2024.

### C. Testing programme

Screening for HIV antibody is available throughout the country and provided at all levels in more than 1300 health facilities and through community-based organisations and virtual HIV self-testing platforms. The number of confirmatory testing laboratories has increased from 197 in 2020 to 251 in 2024, representing a 27% rise. As of October 2024, there are 251 confirmatory laboratories nationwide in all 63 provinces/cities[15]. From 2020 to 2024, the total number of annual HIV screenings remained between 2.7 and 3.5 million, with a stable positivity rate of 0.6% to 0.8% [11] (Figure 6). [16] Testing volume, testing rates, and positivity varied across regions. Ho Chi Minh City was characterised by a high positivity rate and high population testing rate, while the South East region exhibited a relatively high positivity rate with a low population testing rate (Supplementary Figure S2).

**Figure 6.**
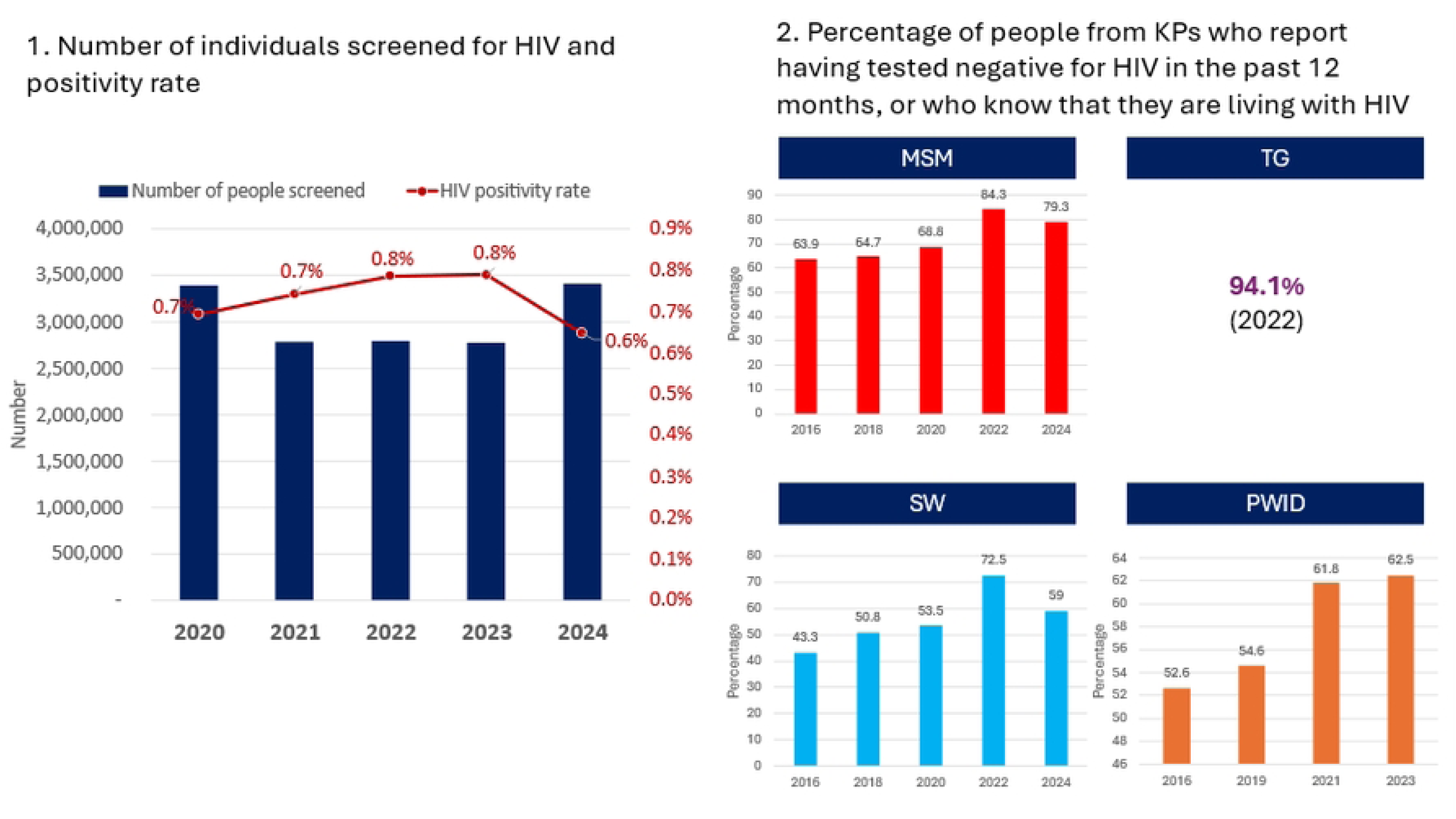
HIV testing among the general population and key populations. KP: key population, MSM: men who have sex with men, PWID: people who inject drugs, SW: sex workers, TG: transgender people.

Among KPs, awareness of HIV status—defined as the proportion who reported testing negative in the past 12 months or knowing they are living with HIV—varied by group: 59.0% among SW, 62.5% among PWID, 79.2% among MSM and 94.1% among TG in 2024[13]. All groups—except TG, for whom trend data are unavailable—already exhibited high HIV status awareness rates in 2016, followed by a further increasing trend in HIV status awareness in recent years (FSW: +15.7%, MSM: +15.4%, PWID: +9.9% since 2016).

### D. Treatment programme

As of 2024, there were 519 outpatient antiretroviral therapy (ART) clinics nationwide, an increase from 451 in 2020. The HIV care cascade highlights the country’s achievement and remaining gaps in diagnosis, treatment initiation, viral load testing, and viral suppression in 2024 (Figure 7) [5]. Of the 234,220 PLHIV who have been diagnosed, 184,222 (79%) were receiving ART. Subnational analysis showed marked variation in ART coverage, ranging from 63.5% in the South-Central region to 81.6% in Ho Chi Minh City (Supplementary Table S1). Among 5,492 PLHIV lost to follow-up, 2,996 (55%) re-engaged in ART in 2024[11].

**Figure 7.**
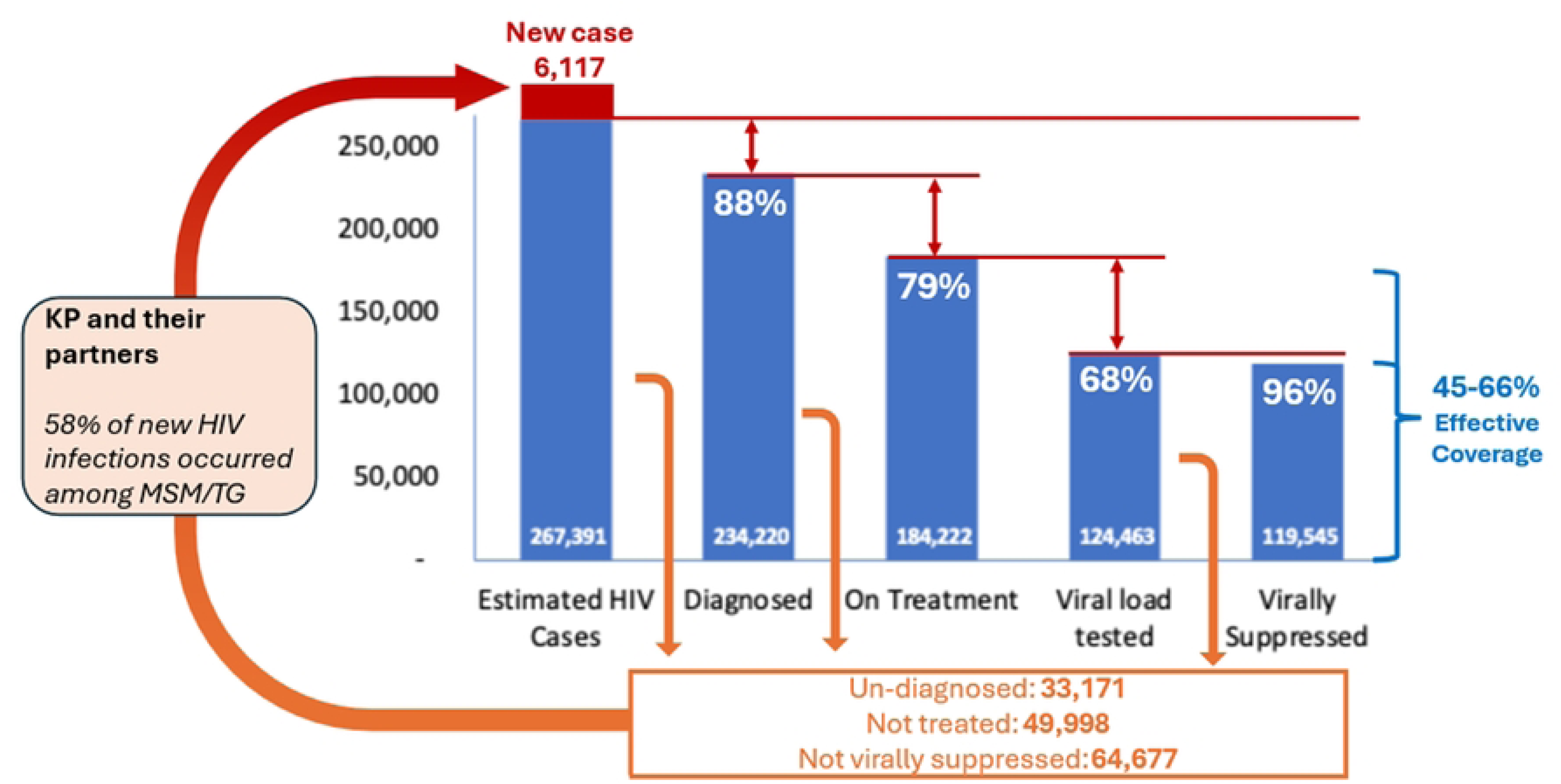
HIV care cascade, 2024 data. KP: key population, MSM: men who have sex with men, TG: transgender people. Effective coverage: Virally suppressed / Estimated HIV cases *100 (1) 119,545 / 267,391*100=45%, (2) (184,222*96%) / 267,391*100=66%

Viral load testing coverage remained at 68%, with testing volumes increasing significantly in recent years: over 120,000 tests were conducted in 2023 and 2024, compared to 102,422 in 2020[11], across 41 facilities in 27 of the 63 provinces (43%). Subnational analysis revealed notable disparities in access to viral load testing across regions. Limited and declining access was observed in the Central Highlands and North Central, whereas coverage remained high in Ho Chi Minh City and the Red River Delta (Supplementary Figure S3). Viral load suppression among those tested was high at 96% (119,545/124,463). Effective coverage—defined as the proportion of all PLHIV who are on ART *and* virally suppressed—ranged from 45% to 66%. Along the care cascade, there are approximately 33,000 (12%) PLHIV who remain undiagnosed, 50,000 (21%) who have been diagnosed but are not receiving treatment, and up to 65,000 (35%) on ART who are not virally suppressed.

## Discussion

Viet Nam has made remarkable progress in responding to HIV over the past decades. A 57% reduction in new HIV infections among adults from 2010 to 2024 is the second-highest in the WHO Western Pacific Region after 58% in New Zealand. Notably, Viet Nam is the only lower-middle-income country that achieved more than a 50% reduction in the Region.

The findings highlight the key drivers of Viet Nam’s successful HIV control: strong political commitment, context-appropriate strategies, robust program implementation across all intervention areas and community engagement. In terms of the prevention programme, Viet Nam has consistently demonstrated exceptional performance in implementing harm reduction services. Considering that not all PWID are opioid-dependent and therefore do not require OST, the reported coverage over the years reflects a remarkably high and sustained level of service. Viet Nam’s first and largest harm reduction initiative - the Preventing HIV in Viet Nam Project - was launched in 2003, when new HIV infections were at their peak (Figure 2) [17]. Following its implementation, new HIV infections among PWID started to decline. The recent data indicate continued high effectiveness of harm reduction programmes, which likely contributed to a significant impact on reducing new HIV infections among PWID. In recent years, as the epidemic has shifted from injecting drug use to male-to-male sexual contact, the country has adopted new prevention strategies for MSM; PrEP was introduced in 2017 through pilot programs at a time when new infections among MSM were on the rise. It was subsequently scaled up with strong political commitment from the Ministry of Health, with a target of achieving 40% coverage among MSM by 2030[3][18]. At the same time, community engagement has played a critical role in reaching populations most in need in Viet Nam [19][20]. These responses reflect Viet Nam’s timely and accurate understanding of the HIV epidemic and its capacity to flexibly implement context-appropriate prevention interventions.

Viet Nam has also demonstrated significant progress in HIV testing strategies. The country’s consistently high testing volume, coupled with recent efforts to further expand and diversify testing service delivery, reflects strong political commitment at the national level. At the same time, Viet Nam has succeeded in maintaining and even improving HIV status awareness among key populations. These achievements indicate the effective implementation of targeted testing approaches focused on groups at the highest risk of HIV acquisition. The country has actively adopted innovative testing approaches over the decades. A notable example of best practice is the successful expansion of HIV testing services through innovative approaches over the decade, including decentralisation of HIV confirmatory testing[21], community-led lay-provider testing and assisted partner notification[22], and web-based HIV self-test distribution with linkage to HIV treatment[23]. This commitment, along with increased testing capacity and a targeted testing strategy, may have contributed to these outcomes in testing programmes.

Viet Nam has adopted and implemented the Treat All strategy [24] and has achieved a high viral load suppression rate, reflecting strong adherence among individuals on ART. On the other hand, it should be noted that of all PLHIV, the percentage of virally suppressed PLHIV is estimated to be 66% at best, and a substantial number of PLHIV were lost along the care cascade. Suboptimal viral load testing coverage is another gap; however, the country has shown efforts to expand testing capacity.

While progress in Viet Nam’s HIV response has been documented [25], our review provides uniquely comprehensive insights into the factors contributing to these achievements based on national programme data. As many countries struggle to achieve their goals amid constrained funding and declining external donor support, learning from established best practices is crucial. Viet Nam’s experience offers valuable insights and may serve as a reference for other low- and middle-income countries facing concentrated HIV epidemics.

Our review also highlights the remaining challenges in Viet Nam, which should be addressed to further accelerate the HIV response in the years leading up to 2030. The plateau in HIV-related mortality, despite high viral suppression among those tested, suggests that late diagnosis, comorbidities in an ageing PLHIV population, and retention gaps may be contributing to ongoing deaths. Prioritising earlier diagnosis, timely ART initiation, and stronger continuity of care will be important to drive mortality down further.

In the context of declining external funding, sustainable financial planning is an urgent issue. While 58 of 63 provinces (92%) have approved a Sustainable HIV Financing Plan and domestic funding accounted for more than 50% of total HIV/AIDS resources during 2021–2024, significant challenges remain in achieving sustainable financing. First, financial resources are insufficient to meet the goals of the National Strategy: from 2021 to 2024, US$432.7 million was mobilised&mdash\semicolon \; only 70&percnt\semicolon \; of the estimated US$619.2 million required for full implementation. Second, available resources are not optimally disbursed: domestic expenditure for 2021–2023 totalled US$91 million\comma \; representing just 54&percnt\semicolon \; of the US$168 million allocated for the same period (unpublished data).

At the same time, strengthened prioritisation coupled with resource allocation optimisation is essential. To further enhance the effectiveness of HIV prevention programs, it is imperative to expand access to PrEP by addressing structural and social barriers, including demand creation [26] and stigma [27]. A substantial proportion of PLHIV remain undiagnosed, diagnosed but not on treatment, or on ART without known viral suppression—populations that may drive ongoing transmission in Viet Nam—underscoring the need to intensify case-finding, enhanced support to ART initiation and continuation, and improved coverage of viral load testing. Additionally, regional disparities were observed in testing and treatment coverage. Enabling equitable access to HIV services across the country is imperative. Ongoing government restructuring may have both transitional risks and opportunities; its impact on HIV service coordination will depend on how decentralisation and integration are implemented.

This study has several limitations. First, it relies on routine program data, which vary in completeness and quality across sites and reporting periods. Inconsistent data entry, missing values, and differences in reporting practices may introduce bias, reduce accuracy, and limit detailed subgroup analyses, affecting the robustness of trend interpretation and regional comparisons. Second, we could not collect sufficient time series for certain program indicators, such as OST coverage, to evaluate long-term implementation, constraining the ability to identify sustained trends and assess the lasting impact of interventions. In particular, we could not evaluate programme implementation in the early 2000s when the decline in new infections began due to limited data availability. Third, the estimates for new HIV infections and PLHIV rely on outputs from the AIDS Epidemic Model (AEM); they are subject to uncertainties inherent to modelled data, including assumptions on key population size estimates, transmission parameters and calibration to surveillance trends.

### Conclusions

Viet Nam’s progress toward ending AIDS has been driven by the country’s strong commitment to context-appropriate strategies and robust program implementation. These achievements provide valuable lessons and can serve as a model for other nations, particularly low- and middle-income countries. Viet Nam will need to address the current challenges and strengthen national efforts to achieve the national target of ending AIDS by 2030.

## Authorship

Conceptualisation: KI, VTTN, FM, PDM, PTTH; Data collection: BHD, DTN, NTMT, DTTL, VTTN; Data analysis: FM, KI, MO; Writing – original draft: MO; Writing – review & editing: KI, VTTN, FM, PDM, PTTH, RPHY, BHD, DTN, NTMT, DTTL; Supervision/Administration: KI, RPHY.

## Acknowledgements

The authors would like to thank the Global Fund to Fight AIDS, Tuberculosis and Malaria for its financial support of the review from which the data for this paper were derived.

The authors also express their sincere appreciation to

- the Viet Nam Administration of Disease Prevention (VADP), Ministry of Health, and other relevant departments, national and regional institutions, provincial Centres for Disease Control, district and commune health facilities, civil society and community-based organizations, in Hanoi, Thanh Hoa, Ninh Binh, Ho Chi Minh City, and Soc Trang for their facilitation and active participation in the review process.
- UNAIDS for jointly conducting the review, with a focus on sustainable financing and procurement, and for its support in reviewing the final report.
- Following individuals for contributing to the HIV program review:

Dr Thuan Van Hoang (WHO Country Office in Viet Nam)

Prof. Abdullah DEMIRKOL (International consultant)

Dr Nathan FORD (WHO Headquarters)

Dr Rachael BAGGALEY (International consultant)

Prof. Peter GODFREY-FAUSSETT (International consultant)

Dr Wayne DIMECH (NRL Australia, WHO Collaborating center) Ms Nguyen Thi Minh Thu (WHO consultant)

## Artificial Intelligence (AI)

Artificial intelligence tools were used solely to improve spelling, grammar, and syntax; no content generation or data analysis was performed.

## Data availability statement

The data used in this study are available from the corresponding author upon reasonable request.

## References

1. United Nations Development Programme Viet Nam. Impact of HIV/AIDS on household vulnerability and poverty in Viet Nam [Internet]. Ha Noi; 2013. Available from: https://www.undp.org/vietnam/publications/impact-hiv/aids-household-vulnerability-and-poverty-viet-nam

2. Coleman M, Akolo C, Mbanusi A, Sithole B, Siberry GK, Schowen R, et al. Integrating HIV and primary healthcare for key populations : community-led models from Vietnam, Nigeria and Eswatini. J Int AIDS Soc. 2025;28.

3. Viet Nam Administration for AIDS Control. National Strategy to End the AIDS Epidemic by 2030. [In Vietnamese]. 2020.

4. Joint United Nations Programme on HIV/AIDS (UNAIDS). AIDSinfo [Internet]. 2025 [cited 2025 Oct 31]. Available from: https://aidsinfo.unaids.org/

5. Ministry of Health Viet Nam. Report on HIV Cascade (2024). 2024.

6. World Health Organization (WHO). Global HIV, Hepatitis and STIs Programmes/Populations [Internet]. [cited 2025 Mar 12]. Available from: https://www.who.int/teams/global-hiv-hepatitis-and-stis-programmes/populations

7. Brown T, Peerapatanapokin W, Siripong N, Puckett R. The AIDS Epidemic Model 2023 for Estimating HIV Trends and Transmission Dynamics in Asian Epidemic Settings. J Acquir Immune Defic Syndr. 2024;95:13–23.

8. World Health Organization (WHO). Consolidated guidelines on person-centred HIV strategic information: strengthening routine data for impact. Geneva; 2022.

9. UNAIDS and Viet Nam Government. the AIDS Epidemic Model. 2025.

10. Ministry of Health Viet Nam. HIV Sentinel Surveillance Plus Behavioural Survey. 2022.

11. Ministry of Health Viet Nam. Viet Nam Administration for AIDS Control. HIV Case Reporting Data (Form C03/C05). 2024.

12. World Health Organization (WHO). Global health sector strategies on, respectively, HIV, viral hepatitis and sexually transmitted infections for the period 2022–2030 [Internet]. 2022. Available from: https://iris.who.int/handle/10665/338901

13. Joint United Nations Programme on HIV/AIDS (UNAIDS). The Key Population Atlas [Internet]. 2025 [cited 2025 Nov 1]. Available from: https://kpatlas.unaids.org/dashboard

14. Ministry of Health Viet Nam. HMED. 2024.

15. Ministry of Health Viet Nam. HIV confirmatory labs, HMED. 2024.

16. United Nations Department of Economic and Social Affairs. World Population Prospects 2024 [Internet]. 2024 [cited 2024 Oct 1]. Available from: https://population.un.org/wpp/

17. World Health Organization. Regional Office for the Western Pacific. Good Practice in Asia : TARGETED HIV PREVENTION FOR INJECTING DRUG USERS AND SEX WORKERS: Viet Nam’s first large-scale national harm reduction initiative [Internet]. 2010. Available from: http://iris.wpro.who.int/handle/10665.1/1388

18. Green KE, Nguyen LH, Phan HTT, Vu BN, Tran MH, Ngo H Van, et al. Prepped for PrEP? Acceptability, continuation and adherence among men who have sex with men and transgender women enrolled as part of Vietnam’s first pre-exposure prophylaxis program. Sex Health. 2021;18:104–15.

19. Sabin LL, Semrau K, Desilva M, Le LTT, Beard JJ, Hamer DH, et al. Effectiveness of community outreach HIV prevention programs in Vietnam: A mixed methods evaluation. BMC Public Health. 2019;19:1–17.

20. Coleman M, Akolo C, Mbanusi A, Sithole B, Siberry GK, Schowen R, et al. Integrating HIV and primary healthcare for key populations : community-led models from Vietnam, Nigeria and Eswatini. J Int AIDS Soc. 2025;28:e70027.

21. Nguyen VTT, Best S, Pham HT, Troung TXL, Hoang TTH, Wilson K, et al. HIV point of care diagnosis: Preventing misdiagnosis experience from a pilot of rapid test algorithm implementation in selected communes in Vietnam. J Int AIDS Soc. 2017;20:67–74.

22. Nguyen VTT, Phan HTT, Kato M, Nguyen QT, Le Ai KA, Vo SH, et al. Community-led HIV testing services including HIV self-testing and assisted partner notification services in Vietnam: lessons from a pilot study in a concentrated epidemic setting. J Int AIDS Soc. 2019;22:40–8.

23. Nguyen VTT, Dunkley Y, Son VH, Choko AT, Huong PTT, Manh PD, et al. Investigating the effectiveness of web-based HIV self-test distribution and linkage to HIV treatment and PrEP among groups at elevated risk of HIV in Viet Nam provinces: a mixed-methods analysis of implementation from pilot to scale-up. J Int AIDS Soc. 2024;27.

24. World Health Organization (WHO). WHO Country Intelligence Tool [Internet]. 2025 [cited 2025 Nov 6]. Available from: https://cfs.hivci.org/country-factsheet.html

25. Chu DT, Vo Truong Nhu N, Tao Y, Le Hoang S. Achievements and challenges in HIV/AIDS control in Vietnam. HIV Med. 2018;19:e75–6.

26. Nguyen MX, Rutstein SE, Hoffman I, Tran H V, Giang LM, Go VF. Low HIV Testing and PrEP Uptake of Adolescent and Young Men who have Sex with Men in Vietnam. AIDS Behav. 2025;29:401–10.

27. Thai TT, Nguyen LT, Hoang HT, Lung NB, Bui TM, Ali M, et al. Homosexuality stigma and HIV risk behaviors among HIV-negative men who have sex with men in Vietnam ABSTRACT. AIDS Care [Internet]. 2025;37:1058–66. Available from: 10.1080/09540121.2025.2476636

